# Long-Term Persistence of IgG Antibodies in SARS-CoV Infected Healthcare Workers

**DOI:** 10.1101/2020.02.12.20021386

**Authors:** Xiaoqin Guo, Zhongmin Guo, Chaohui Duan, Zeliang Chen, Guoling Wang, Yi Lu, Mengfeng Li, Jiahai Lu

**Author notes:** Corresponding author. for Jiahai Lu. the authors contributed equally to this work (Xiaoqin Guo, Zhongmin Guo, Chaohui Duan, Zeliang Chen).

## Abstract

**BACKGROUND:** The ongoing worldwide outbreak of the 2019-nCoV is markedly similar to the severe acute respiratory syndrome (SARS) outbreak 17 years ago. During the 2002-2003 SARS outbreak, healthcare workers formed a special population of patients. Although virus-specific IgG play important roles in virus neutralization and prevention against future infection, limited information is available regarding the long term persistence of IgG after infection with SARS-like coronavirus.

**METHODS:** A long-term prospective cohort study followed 34 SARS-CoV-infected healthcare workers from a hospital with clustered infected cases during the 2002-2003 SARS outbreak in Guangzhou, China, with a 13-year follow-up. Serum samples were collected annually from 2003-2015. Twenty SARS-CoV-infected and 40 non-infected healthcare workers were enrolled in 2015, and their serum samples were collected. All sera were tested for IgG antibodies with ELISA using whole virus and a recombinant nucleocapsid protein of SARS-CoV, as a diagnostic antigen.

**RESULTS:** Anti SARS-CoV IgG was found to persist for up to 12 years. IgG titers typically peaked in 2004, declining rapidly from 2004-2006, and then continued to decline at a slower rate. IgG titers in SARS-CoV-infected healthcare workers remained at a significantly high level until 2015. Patients treated with corticosteroids at the time of infection were found to have lower IgG titers than those without.

**CONCLUSIONS:** IgG antibodies against SARS-CoV can persist for at least 12 years. The presence of SARS-CoV IgG might provide protection against SARS-CoV and other betacoronavirus. This study provides valuable information regarding humoral immune responses against SARS-CoV and the 2019-nCoV.

## 1. INTRODUCTION

On December 31, 2019, an outbreak of coronavirus causing infections in Wuhan, China, was officially reported. The outbreak rapidly spread to several provinces of China and other countries. Currently, the ongoing outbreak in China caused by a new coronavirus, named 2019-nCoV by WHO, is becoming a global crisis. Notably, the 2019-nCoV is highly similar to the severe acute respiratory syndrome coronavirus (SARS-CoV), which is characterized by high mortality and infectivity. Therefore, information on SARS-CoV will be valuable for the prevention and control of the current outbreak.

SARS was first confirmed in Guangdong, China in 2002-2003, and subsequently became a pandemic, spreading rapidly to 31 countries and regions, causing panic and a global public health crisis.^1-3^ In total, 8,422 SARS cases were reported to the World Health Organization (WHO) with a case fatality rate of 10.88% (916/8,422); of these total cases, 5,327 cases and 349 deaths were reported from 29 provinces of China. Guangdong was among the most affected provinces, with 1,511 reported cases, 22.90% (346/1,511) of which occurred in healthcare workers.^3-5^ During emergency disease control, frontline healthcare workers, including physicians, nurses, and other healthcare workers, who were exposed to the virus while treating and caring for SARS patients in the early stages of the outbreaks, were mainly affected. During the later stages of the outbreak, although precautions to prevent the infection were enhanced, infection of healthcare workers still occurred.^6,7^ No natural SARS-CoV infections have been identified since the last confirmed case in May 2004.^8^ The reasons why SARS caused a transient epidemic and was experienced as an isolated event, remain unelucidated.^9^ There are concerns that SARS-CoV will re-emerge in the future; consequently, studies are underway to determine the mechanisms through which SARS-CoV interacts with the host immune system, crosses host barriers, and other related aspects.

SARS-CoV was initially identified as having occurred through interspecies transmission from small carnivores (e.g., palm civets and raccoon dogs) to humans in 2003.^10^ Recently, studies indicate that SARS-CoV may have originated from bats. Bat coronavirus isolates have been shown to be able to recognize both human and bat ACE2 receptors.^11^ Coronaviruses can exploit numerous cellular receptors and potentially cause various illnesses such as respiratory and gastrointestinal diseases.^12^ Particularly, the spike protein is an important viral determinant of virus-receptor interaction, interspecies transmission, and host tropism, and its mutation can facilitate cross-species infection and human-to-human transmission.^13^ Another novel coronavirus similar to SARS-CoV, the middle-east respiratory syndrome coronavirus (MERS-CoV), emerged in the Middle East in 2012. MERS-CoV has been shown to be transmissible from camels to humans, highlighting the ongoing threat posed by SARS-CoV or other SARS-like viruses.^14,15^

SARS-CoV IgG antibodies have neutralization activities and provide protection against infections. Presence of IgG antibodies against virus and nucleocapsid protein represents protective immune responses. While some previous studies reported longitudinal profiles of SARS IgG antibodies for up to 5 years,^16-20^ there has been a lack of knowledge on humoral immune responses for longer durations. In our current study, we conducted a 13-year follow-up study on a cohort of SARS-CoV-infected healthcare workers for analyzing the presence and persistence of antibodies against SARS-CoV.

## 2. METHODS

### 2.1 STUDY SITE AND PARTICIPANT ENROLLMENT

The study site was a teaching hospital and the main center for treating clustered SARS cases in 2002-2003 in Guangzhou, China. The hospital employed 4,797 staff, possessed 2,140 annual open beds, and treated over 3.2 million patients annually. During the outbreak (from February to March, 2003), a total of 95 healthcare workers, out of all the staff in this hospital, were diagnosed with SARS according to the diagnostic criteria promulgated by the Health Bureau of Guangdong (published on February 3, 2003), and 34 of these cases were enrolled in this study. The majority of the participants were aged between 20 and 30, in 2003, and 94.11% (32/34) of them were females (Table S1). The enrollment criteria included clinical diagnosis of SARS-CoV infection in a healthcare worker treated the above mentioned hospital, the date of onset being prior to April 16, 2003, and an estimated long-term follow-up of more than 9 years.

The sera used in this study is derived sample of the annual physical examination of the SARS-CoV infected healthcare workers. Antibody surveillance is a part of the examination. The annual physical examination has been reviewed and supported by the hospital ethics review committee. Written consent was signed at the first examination.

### 2.2 SAMPLE COLLECTION

A total of 362 serum samples was collected. This included samples collected at the time of hospital admission in 2003, from all study participants, and yearly sample collection, when the participants attended their annual phyical examinations (April to June from 2004 through 2015). Eighty samples were missing in the follow-up (Table S2): there was an interruption of sample collection in 2012 or 2013 due to an unexpected absence of the investigators in charge of sample collections; samples of 6 subjects were not collected in 2014 (Patient No. 29 - 34) and 2015 (Patient No. 26 - 28, 31-33) respectively, for failing to arrange a physical examination or due to resignation. In addition, serum samples from another 20 subjects, from the 95 SARS cases, were collected in 2015 (Table S3). Another 40 non-SARS infected healthcare workers from the same hospital were enrolled and serum samples were collected, as the control group, to verify the end result data of the 34 SARS-CoV-infected healthcare cohort in 2015 (Table S4).

### 2.3 DETECTION AND QUANTIFICATION OF IgG ANTIBODIES

Serum IgG antibodies were detected with a SARS-CoV IgG antibody enzyme-linked immunosorbent assay (ELISA) Kit (BGI-GBI Biotech Co. Ltd., Beijing, China), which used the whole SARS-CoV as the detection antigen. Briefly, the serum samples were added at a 1:10 diluion and incubated in pre-coated plates. After the processes of washing, reacting with HRP-labeled goat anti-human IgG, color developing, and stopping the reaction, the absorbance was measured at 450 nm using a plate reader. The IgG antibodies against nucleocapsid protein were detected with a recombinant SARS-CoV N199 antigen assay developed in our laboratory as described previously^21^. Indirect ELISA was performed to detect and quantify antibodies (at 1:10 dilution) against SARS-CoV N199 in the serum samples, as previously described by Guo et al^21^.

### 2.4 STATISTICAL ANALYSIS

Boxplots and heatmaps were used to show the temporal trends and variations in the antibody levels. Non-linear exponential decay models were used to estimate the average decay rates of the antibody level. A non-parametric Mann Whitney U test was used to compare the IgG values of samples from SARS-CoV-infected healthcare workers with those from the control in 2015. The test was also used to compare the anti-SARS-CoV IgG titers from SARS-CoV-infected healthcare workers who were treated with or without corticosteroids in 2003. We used the Kappa test to estimate the consistence of the results of a positive rate for both methods in the samples of the SARS-CoV-infected healthcare worker group from 2003 to 2005 and non-infected healthcare worker group in 2015. Missing values were replaced with estimated values from exponential decay models. Additionally, we conducted a comparison for the decay trend of the data with and without the missing value replacements.

## 3. RESULTS

### 3.1 PROFILES OF IgG AGAINST SARS-CoV FROM 2003 TO 2015

ELISA detection results showed that the IgG levels peaked in 2004, declined quickly from 2004 to 2006, and further decreased at a slower rate to a mean OD_450nm_ value of 0.286 (against whole virus) and 0.330 (against N199) in 2015 (Figure 1, A and B). IgG levels against SARS-CoV varied between individuals, with the OD_450nm_ range from 1.162 to 0.48 for IgG against whole virus, and 2.374 to 0.581 for IgG against N199. Notably, two healthcare workers, who were diagnosed as SARS-CoV-infected patients, were negative for IgG against both the whole virus and N199, over the entire study period (2003 – 2015), suggesting a possibility of misdiagnosis during the initial admission in 2003; therefore, they were excluded from further statistical analyses (Figure 1, C and D).The IgG levels against whole virus in 4 SARS infected healthcare workers increased notably after 2004, two of which also showed an increase in IgG antibodies against N199, while their levels in other SARS infected healthcare workers decreased continuously during the following years (Figure 1,C, and D).

**Figure 1.**
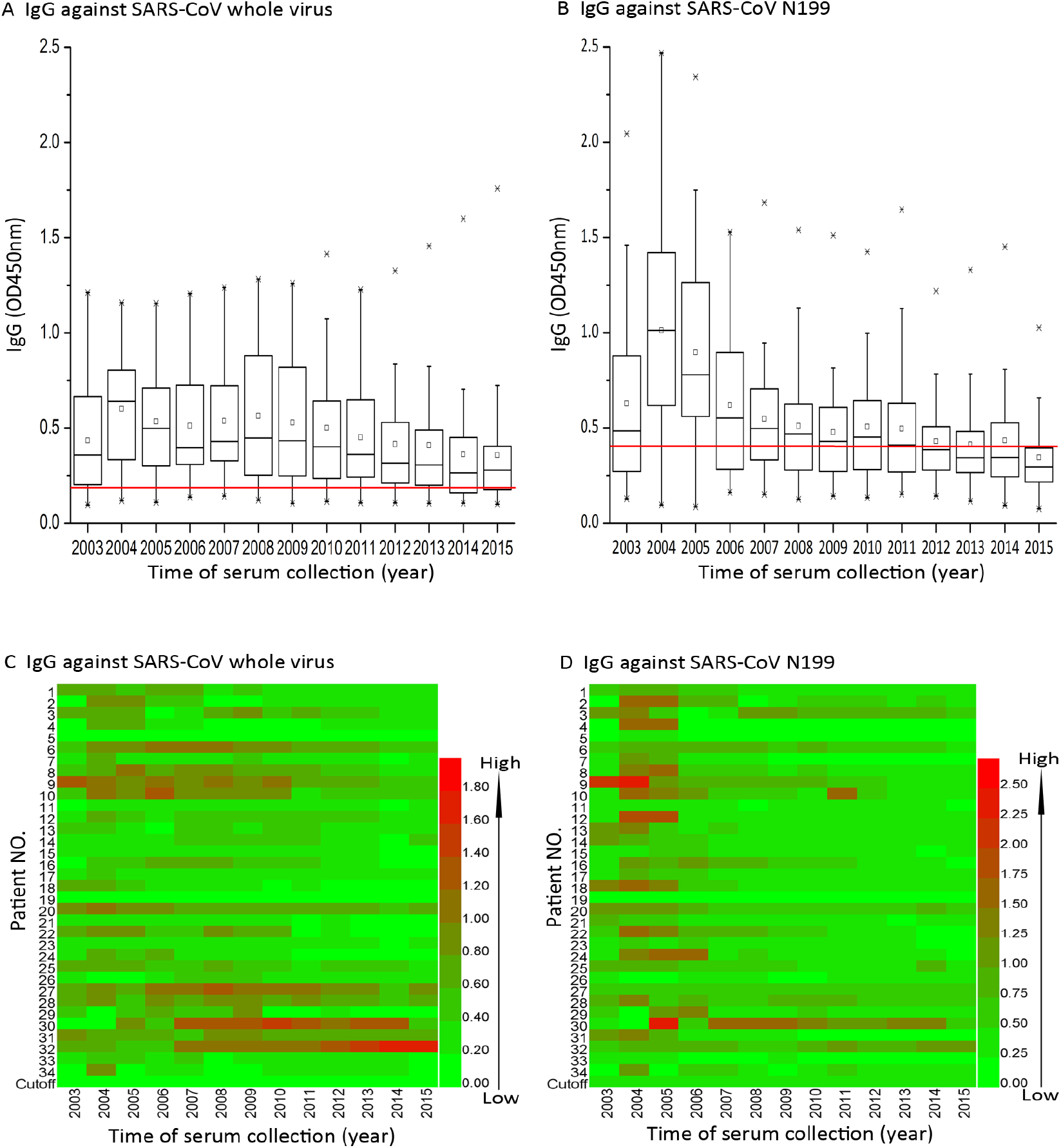
Temporal changes in IgG antibodies against SARS-CoV whole virus and N199 in SARS-Cov-infected healthcare workers. (A) The levels of IgG antibodies using SARS-CoV whole virus as antigen for ELISA detection. The boxplot displays the IgG titers of the 25th, 50^th^, and 75th percentiles. The whiskers extend to the most extreme data point <1.5 times the inter-quartile range. The cutoff value (red lines) was defined at 0.19 according to the manufacturer’s instructions for the SARS-CoV whole virus ELISA kit. Eighty samples lost during follow up were estimated for their reading values using exponential regression of curve estimation method (The same as below). (B) Levels of IgG antibodies against SARS-CoV N199. The cutoff value (red lines) was calculated as follows: cutoff = mean of OD_450nm_ from 52 general population serum samples +3×standard deviations, which gave a value of 0.40. (C) The dynamics of IgG against SARS-CoV whole virus of each individual, from 2003 to 2015, illustrated by heatmap (HemI 1.0, Heatmap Illustrator).^26^ (D) The dynamics of IgG against SARS-CoV N199 of each individual from 2003 to 2015.

In the 13-year follow-up cohort, 80 out of 442 (18.10%) serum samples were lost. We used the exponential regression of curve estimation method to estimate the missing values (Figure 1). Next, we adopted the exponential decay models to define antibody decay, and the results indicated that the antibody titers exhibited an overall declining trend in IgG against both whole virus and N199 (Figure 2 A and B). A similar decay trend inf IgG titers was found, although the missing values were not replaced in the model (Figures S1 and S2). Furthermore, the proportion of study participants with IgG titer against whole virus (> cutoff value) was 81.25% (26/32) in 2003, peaked at 100.00% (32/32) in 2007, and decreased to 69.23% (18/26) in 2015 (Figure 2C). For IgG antibody against N199, the initial positive (higher than the cutoff value) was 59.38% (19/32) in 2003, peaked at 87.50% (28/32) in 2005, and finally decreased to 19.23% (5/26) in 2015 (Figure2D). The proportion of both the IgGs exhibited a reduced trend, and the higher level (>2 cutoff value) declined annually until it reached 26.92 % (IgG against whole virus) and 0.00% (IgG against N199) in 2015 (Figure 2, C and D). Two healthcare workers, patient No. 11 and No. 26, who had previously been diagnosed with SARS-CoV infection, were identified not to have detectable levels of IgG against N199 over the entire study period (2003-2015). Anti-N199 IgG titers for patient No. 33 were also negative for all the samples collected except the ones collected in 2005. The IgG titer against the whole virus in these patients were weakly positive.

**Figure 2.**
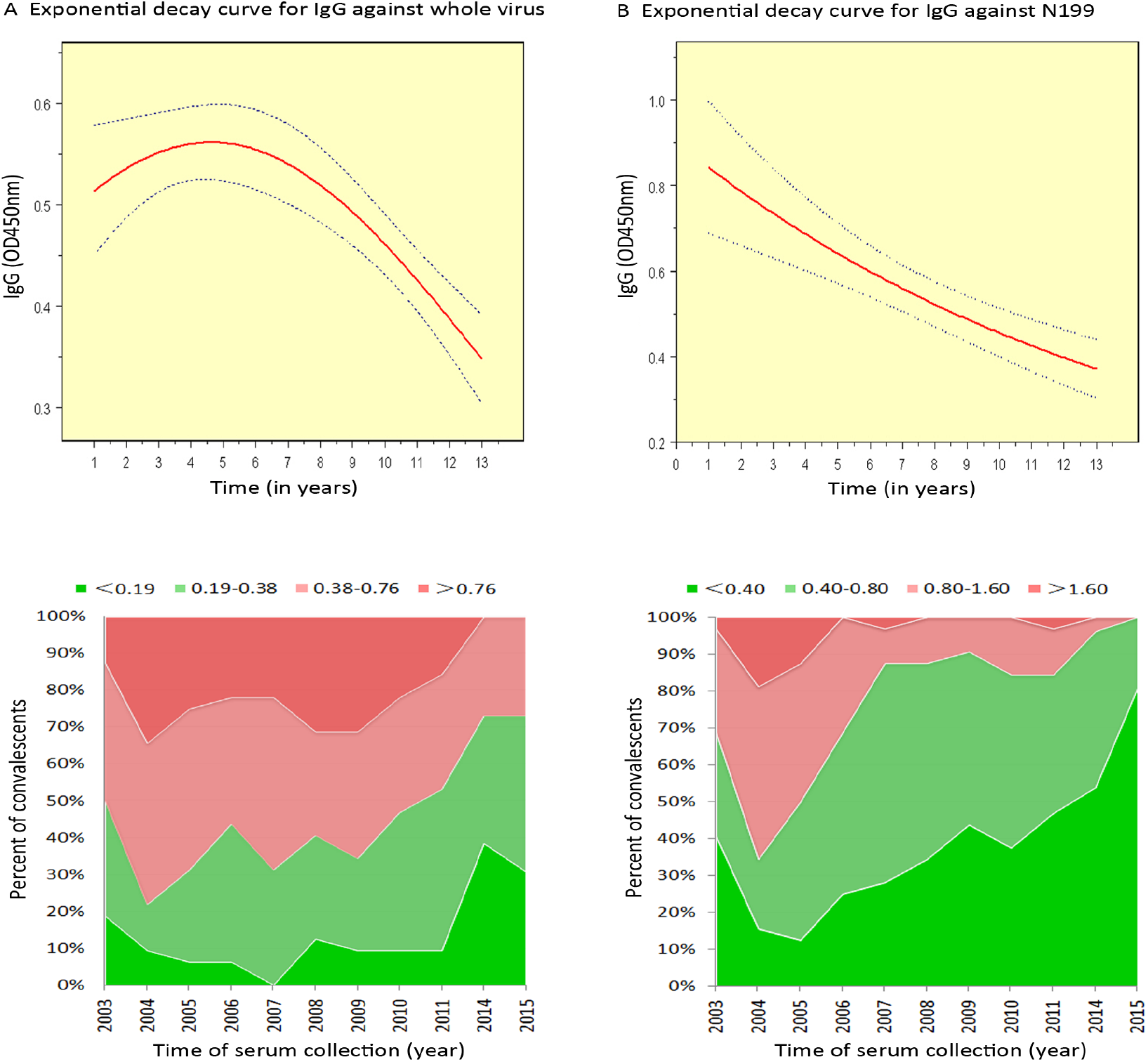
Population profiles of IgG against SARS-CoV whole virus and N199 from 2003 to 2015. (A) Predicted mean (red line) with 95% confidence limits (blue lines) of IgG against whole virus in the cohort from 2003 to 2015. Eighty specimens lost during follow up were estimated for the missing values using the exponential regression of curve estimation method (The same as Panel B). (B) Predicted mean with 95% confidence limits of IgG against N199 in the cohort from 2003 to 2015. (C) Proportions of different ranges of IgG against SARS-CoV whole virus in the SARS-CoV-infected healthcare workers. (D) Proportions of different ranges of IgG against SARS-CoV N199 in SARS-CoV-infected healthcare workers

### 3.2 DIFFERENCES IN ANTI-SARS-CoV IgG TITERS OF DIFFERENT SARS-CoV-INFECTED GROUPS

To analyze the general IgG distributions, subjects were divided into 2 groups: group 1 (n=46), including SARS-CoV-infected healthcare workers enrolled in the original cohort (n=26) and an additional group of SARS-CoV-infected healthcare workers who were enrolled in 2015 (n=20), and group 2 (n=40) of non-infected healthcare worker, selected as the control. The IgG against whole virus was above the cutoff value, and the IgG titers were significantly higher in group 1 than in group 2 (p < 0.001; Figure 3A). A similar trend was observed for IgG against N199 titers, as group 1 had significantly higher IgG titers than group 2 (p=0.034; Figure 3B). Moreover, in group 1 the positive rate of IgG binding to whole virus and N199 was 58.70% and 28.26% with statiscical significance (p < 0.005), respectively.

**Figure 3.**
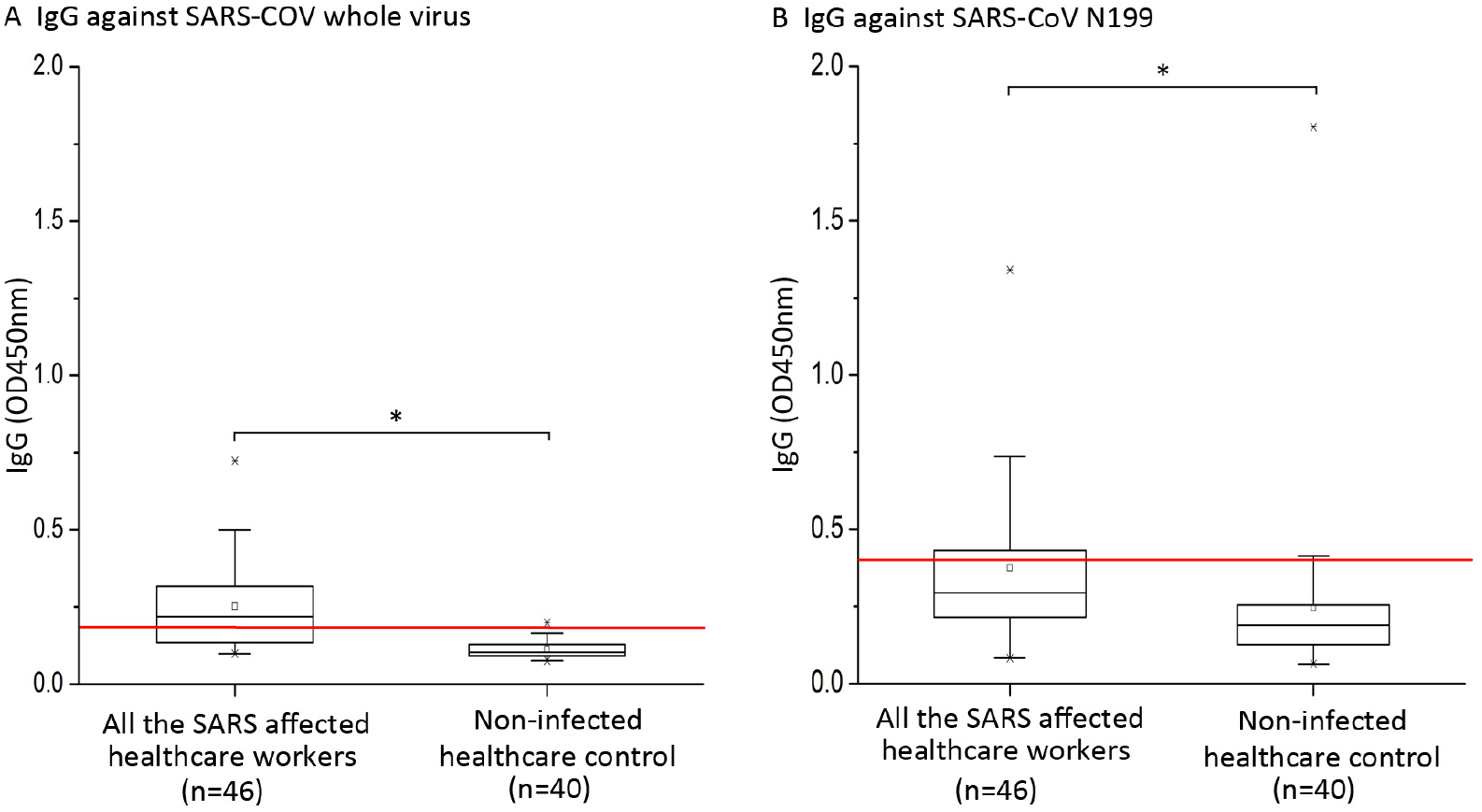
Distributions and differences of IgG in SARS-infected and non-infected healthcare workers (control) (A) The distributions and differences of IgG antibodies against SARS-CoV whole virus between all SARS-CoV-infected (n=46) and non-infected healthcare workers (control) enrolled in 2015 (n=40). The boxplot displays the IgG titers of the 25th, 50th, and 75th percentiles. The whiskers extend to the most extreme data point <1.5 times the inter-quartile range. The red line represents the cutoff value (0.19). (B) The distributions and differences of IgG antibodies against SARS-CoV N199 between all the SARS-CoV-infected (n=46) and non-infected healthcare worker controls enrolled in 2015 (n=40). The red line is the cutoff value (0.40). *P* value of < 0.05 (two-tai) is considered to be statistically significant (*).

Kappa test was used to estimate the consistency between the methods for testing IgG. The consistency test was mainly focused on the period from 2003-2005, as the IgG titers remained stable during this period. Over this period, only 10 of the 102 cases showed inconsistent results between the two detection methods, and the Kappa coefficient was 0.712, suggesting a good general consistence between the results obtained through the whole virus and N199 ELISA methods. Similarly, in the control group, only 5 of the 40 cases displayed inconsistent results, and the Kappa coefficient was −0.42, also indicating a general data consistency between the two methods (Table S5).

### 3.3 THE EFFECT OF CORTICOSTEROID TREATMENT ON IgG ANTIBODY RESPONSES

During the outbreak, some SARS-CoV-infected healthcare workers received corticosteroid treatment, while others did not. The cohort was divided into two groups to assess the effect of corticosteroid use on IgG titers in 2003. Of the 32 SARS-CoV-infected patients (with 2 cases excluded as suspected to have been misdiagnosed), medical records were available for 27 patients, and the records showed that 17 of these patients were, and the rest were not, treated with corticosteroids during the initial care for SARS in the 2002-2003 outbreak (Table S6). In SARS patients with corticosteroids treatment and those not treated with corticosteroids, the IgG against the whole virus was 0.35 ± 0.24 and 0.58 ± 0.29, respectively (p=0.03; Figure 4A), and the IgG against N199 was 0.56 ± 0.40 and 0.72 ± 0.51, respectively. Although IgG against N199 of patients not treated with corticosteroids were higher than those treated with corticosteroids, the difference was not statistically significant (Figure 4B; p=0.17).

**Figure 4.**
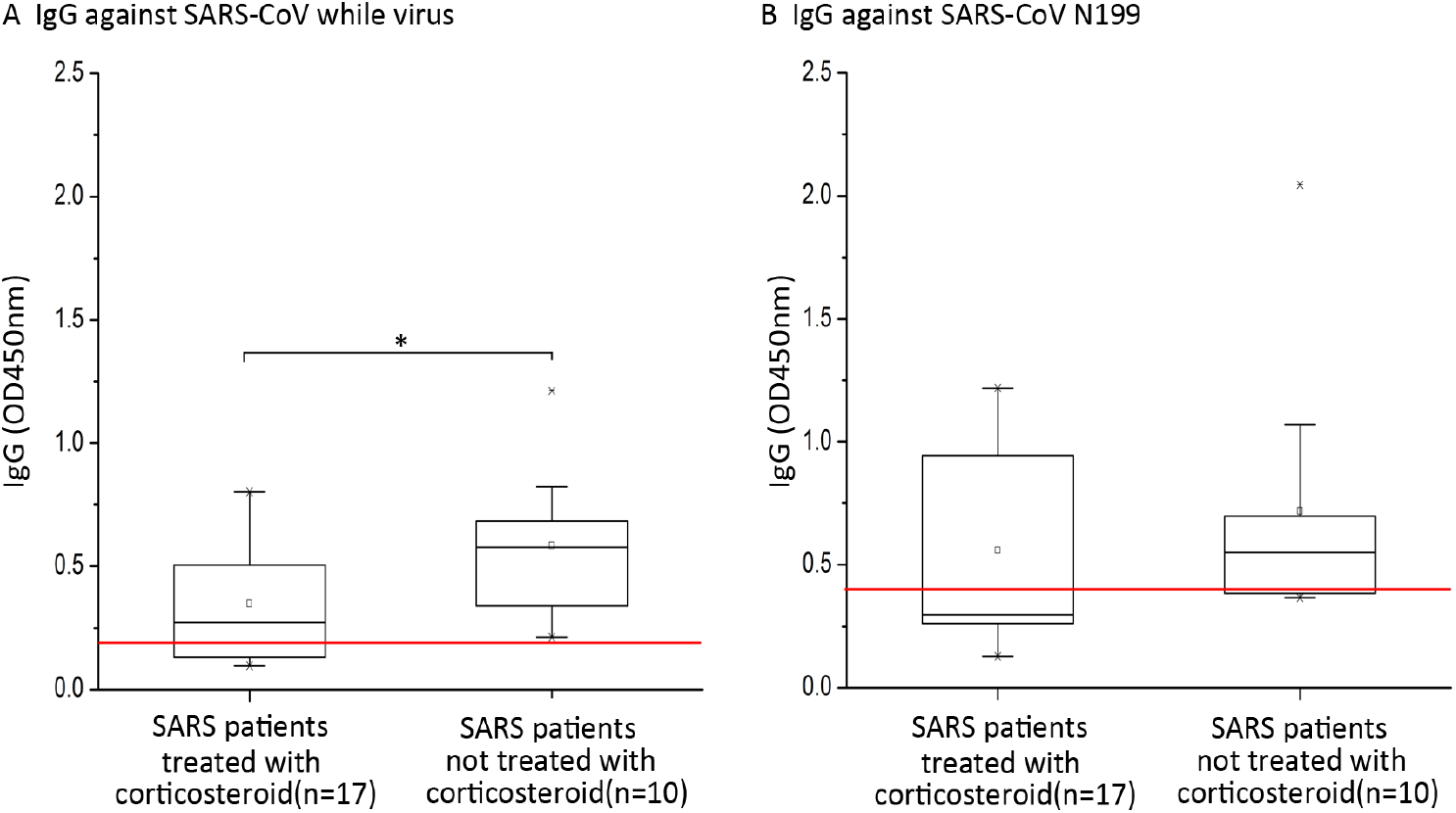
Differences of IgG titers between SARS patients treated with or without corticosteroids. Difference in IgG titers against SARS-CoV whole virus (A) or SARS-CoV N199 antigen (B) between SARS-infected healthcare workers treated with or without corticosteroids. The boxplot displays the IgG titers of the 25th, 50th, and 75th percentiles. The whiskers extend to the most extreme data point <1.5 times the inter-quartile range. The red line represents the cutoff value (0.19, Figure 4A; 0.40, Figure 4B), *P* value of < 0.05 (two-tai) is considered to be statistically significant (*).

## 4. DISCUSSIONS

SARS was first confirmed in Guangdong, China in 2003, and subsequently became a pandemic as it spread rapidly to other countries and regions. Cases from Guangdong accounted for nearly 20% of reported cases and approximately 17% of reported healthcare worker cases worldwide. Healthcare workers, in particular, were exposed to an extremely high risk in combating SARS due to limited knowledge of the etiology and other natures of SARS outbreak. Presently, 17 years after the last SARS outbreak, a new SARS-CoV-like virus, 2019-nCoV, a new member of the betacoronavirus, is causing large scale outbreak in China. Till January 21, 2010, 440 confirmed cases have been reported in China, and several cases have been reported outside China, including the USA, Japan, and Thailand. Unfortunately, a large cluster of healthcare worker infections was officially reported on January 20, 2020 at an early stage of this outbreak. Since information regarding this new betacoronavirus outbreak remains largely unknown, evidence and knowledge from SARS-CoV may provide important clues, aiding our responses to the outbreak.

IgG antibodies are proven to be important for virus neutralization and immunological protection. As the first sampling of sera was performed during the early stage of SARS, it was not possible to determine the maximum antibody titer, as this would be expected to occur approximately 4 months after the onset of the outbreak.^19^ Knowledge of the persistence and level of antibodies against SARS-CoV, particularly neutralizing antibodies, is important for evaluating the immune protection duration and vaccine efficacy. Neutralizing antibodies are normally tested by viral neutralization tests; however, as manipulation involving live SARS-CoV is prohibited in China in the interest of preventing laboratory-acquired SARS infection and potential outbreaks, an alternative approach was employed to detect specific IgG antibodies using SARS-CoV whole virus as capture antigens, which has been suggested in previous studies to correlate with neutralizing activity^16^. Moreover, the SARS-CoV N199 antigen was used to detect its specific antibody, which has been demonstrated to be specific in clinical diagnosis and SARS-CoV reservoir screening.^21^ We therefore adopted the testing of IgG against N199, combined with the titer analysis of IgG against the whole virus, to comprehensively evaluate the SARS antibody duration.

We found that the SARS-CoV-infected healthcare workers who were treated with corticosteroids, such as prednisone and methylprednisolone, exhibited lower IgG titers during the early stages of the disease than those who were not treated with corticosteroids (although not significantly for IgG against N199). However, no differences were found between the two groups after 2003, suggesting that corticosteroids may suppress the early production of SARS-CoV IgG antibodies.

In our cohort, we found that two healthcare workers might have been misdiagnosed with SARS, as the IgG antibodies against both the whole virus and N199 antigen was persistently lower than the cutoff value, for these two patients. Notably, all the cases enrolled in this study were admitted to the hospital between February and March, 2003, and that all the diagnoses made before WHO’s announcement that SARS-CoV was the causative agent of SARS on April 16, 2003^1^, were based on the epidemiological features, clinical symptoms, and regular laboratory blood tests, because serological and etiological diagnostic methods were not available. Specifically, atypical pneumonia, as it was commonly known prior to adoption of the SARS nomenclature, could also be caused by other pathogens.^22^ Based on the results of our current 13-year serological follow-up study, it is possible that the two healthcare workers might have been misdiagnosed. Thus, it warrants further investigation to clarify whether these two subjects were non-responders who did not develop anti-SARS-CoV antibodies at all or simply represented misdiagnosed cases. In the latter scenario, the number of SARS cases in the early stage of 2002-2003 SARS outbreak may need to be reassessed. Presently, for the 2019-nCoV outbreak, nucleic acid-based assay and particular real time PCR, are the mainstream methods for etiological diagnosis. However, the sensitivity and specificity of these assays need careful evaluation, because any false positive or negative results might be harmful to the subjects.

During the SARS outbreak, the typical SARS patients mainly presented with antibodies positive for SARS-CoV, fever, and symptoms of respiratory tract infections. Additionally, there was a small group of asymptomatic SARS-CoV-nfected subjects, who only presented with antibodies positive for SARS-CoV, but showed an absence of clinical symptoms.^23-25^. In this study, we found that four subjects had an abnormal increase in the titers of SARS-CoV antibodies during the follow-up. In contrast, the titers of SARS-CoV antibodies in most of the other healthcare workers appeared to decrease slowly. It is unclear if these four healthcare workers were reinfected with SARS-CoV or other coronaviruses with no signs or symptoms. Thus, further investigation is needed to clarify whether these healthcare workers elicited IgG response by SARS-CoV or other coronaviruses. In the former situation, the novel mechanism of IgG generation without the clinical SARS symptoms needs to be elucidated.

Although IgG against SARS-CoV declined from 2003 to 2015 gradually, IgG levels in SARS-CoV-infected healthcare workers were significantly higher than those in the non-infected healthcare workers control group in 2015. The infected cases enrolled into the study in 2015, exhibited detectable IgG against whole virus, and the positive rate was ∼60%. For IgG against N199, the positive rate was ∼30% in 2015. Moreover, results of the exponential decay curve revealed that IgG antibodies against the whole virus were positive throughout the 13-year follow-up, while IgG antibodies against N199 were negative only in 2015.

Collectively, based on our results, we can infer that the IgG against SARS-CoV can persist at least for 12 years. Presently, we are unaware whether the persistent IgG antibodies possess virus neutralization activities and can provide complete protection against SARS-CoV infection or cross protection against the new betacoronavirus, 2019-nCoV. From previous studies, we predicted that the presence of IgG antibodies could at least provide a partial protection against coronavirus infection. These data will be valuable for better understanding the immune mechanisms and vaccine development strategies against SARS-CoV and 2019-nCoV, therefore being helpful for responses against the ongoing coronavirus outbreak.

## Data Availability

Data are available by requirement from corresponding authors.

## ACKNOWLEDGEMENT

We thanks collaborators participating in sample collection during the cohort study. This work is supported by Guangdong Province Key Area R & D Plan Project (2018B020241002).

## AUTHORS’ CONTRIBUTION

J. Lu designed the study, X. Guo, Z. Guo performed the ELISA tests, Z. Duan collected samples, Z. Chen analyzed the data and revised the manuscript. G. Wang, Y. Lu and M. Li participated in data analysis.

## Supplementary Appendix

**Figure S1.**
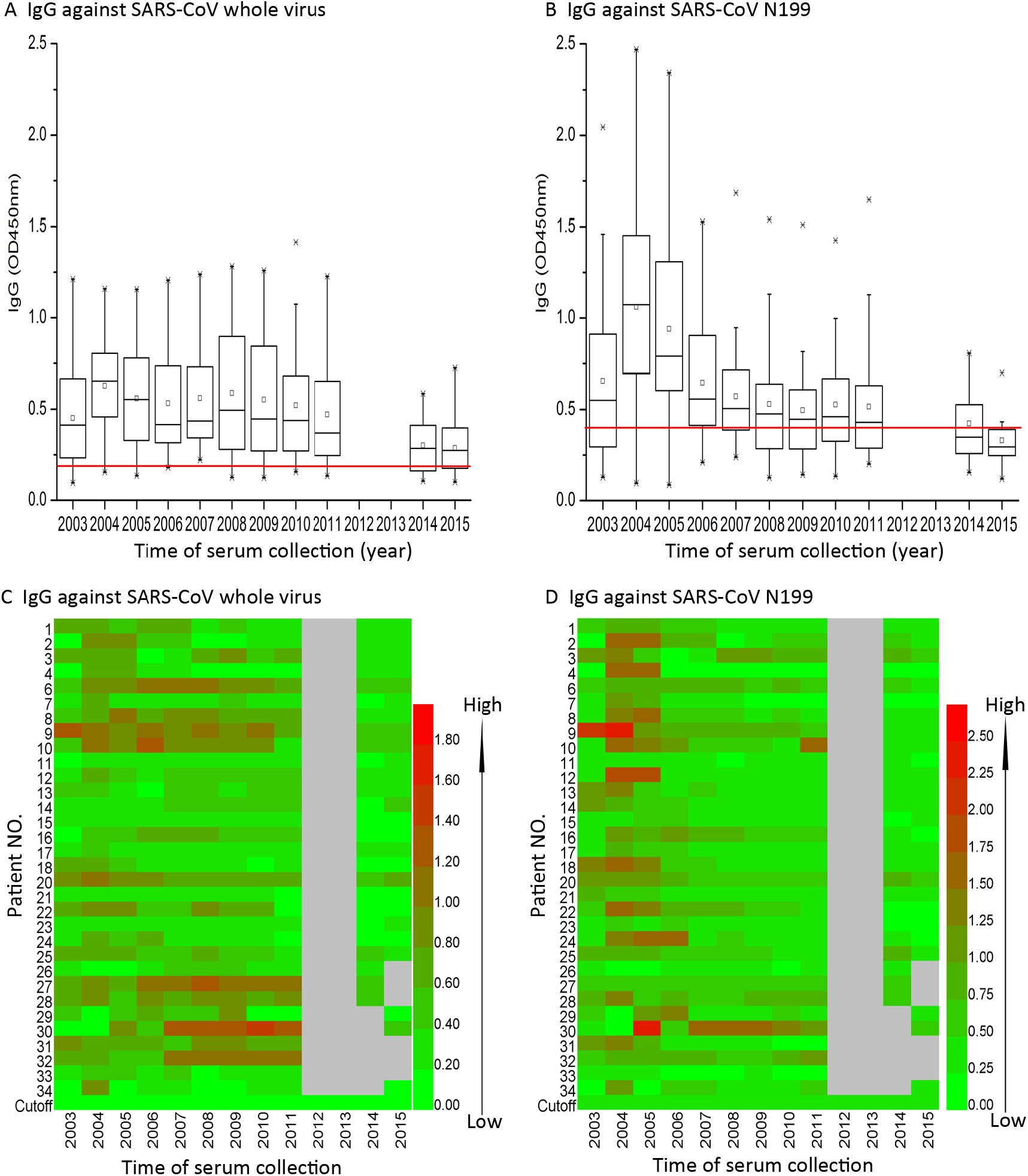
Temporal changes in IgG antibodies against SARS-CoV whole virus and N199 in the SARS-infected healthcare workers (80 predicted missing values excluded) (A) Level of IgG antibodies against SARS-CoV whole virus. The boxplot displays the IgG tiers of the 25th, 50th, and 75th percentiles. The whiskers extend to the most extreme data point <1.5 times the inter-quartile range. The cutoff value (red line) was defined as a value of 0.19 according to the manufacturer’s instruction of the SARS-CoV whole virus ELISA kit. (B) Levels of IgG antibodies against SARS-CoV N199. The cutoff value (red line) was calculated as follows: cutoff = mean of OD_450nm_ from 52 general population serum samples+3X standard deviations, which gave a value of 0.40. (C and D) dynamics of IgG titers against SARS-CoV whole virus (C) or SARS-CoV N199 (D) of each individual from 2003 to 2015 by the method described above. Gray blanks represent the serum samples lost.

**Figure S2.**
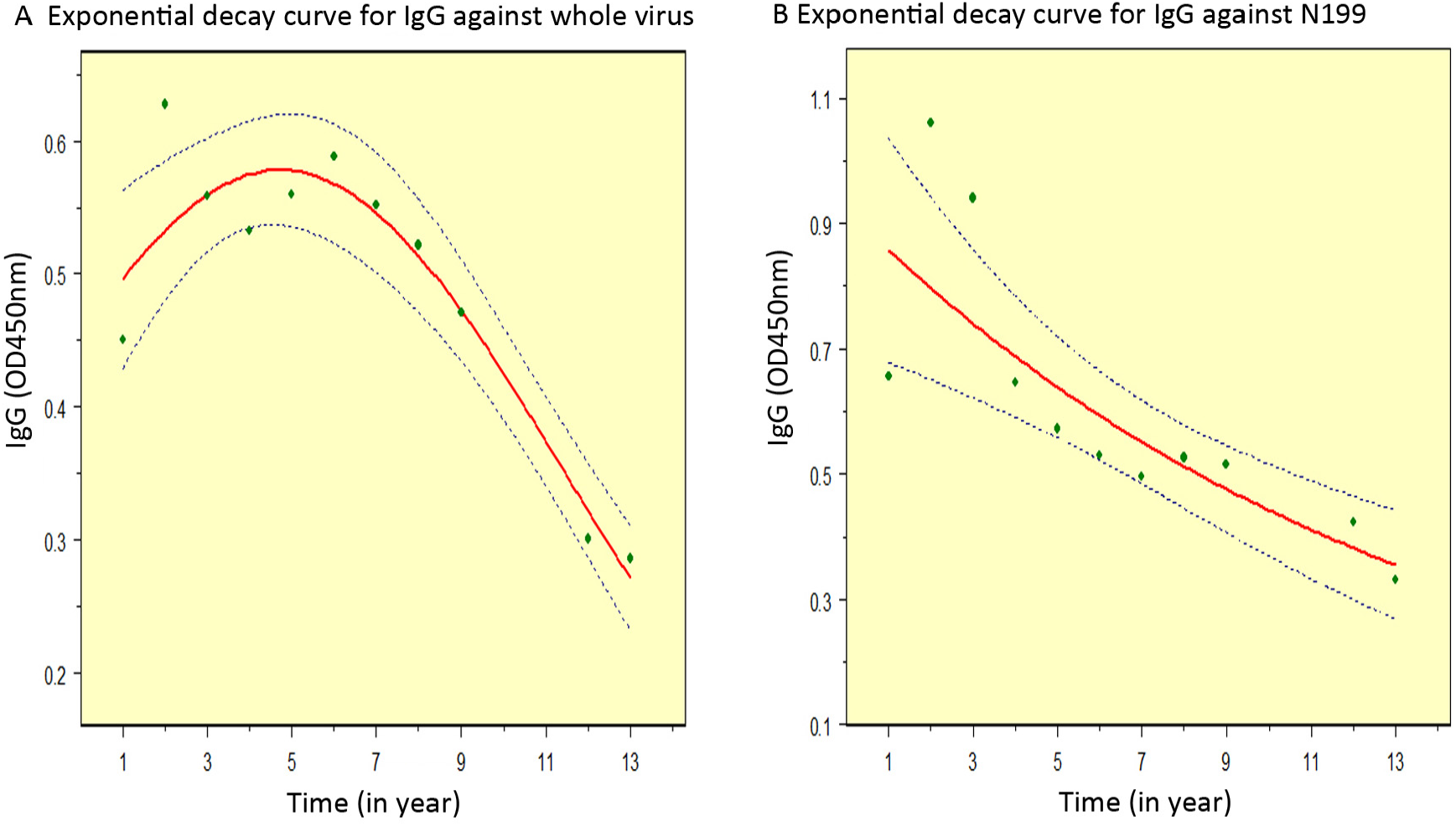
Predicted means of titers of IgG against SARS-CoV whole virus and N199 during 2003 - 2015 (80 predicted missing values excluded) Predicted means (red lines) and actual values (green solid circles) with 95% confidence limits (blue lines) of IgG titers against the whole virus (A) and N199 antigen (B) in the cohort from 2003 to 2015.

